# Evaluating the autoantibody reactome in giant cell arteritis

**DOI:** 10.64898/2026.07.02.26357160

**Authors:** Matthew Porteous, Robert T. Maughan, Louise Sorensen, Michal Zulcinski, Aamir Aslam, Sarah L. Mackie, Charis Pericleous, James Tomlinson, Raashid A. Luqmani, Matthew C. Pickering, Ann W. Morgan, James E. Peters

## Abstract

**Objective:** To determine whether autoantibodies are present in giant cell arteritis (GCA) using a high-dimensional autoantibody array.

**Methods:** Serum was collected from patients with GCA (n=20), other related vascular inflammatory diseases (Takayasu arteritis n=12, IgG4-RD n=5, Behcet’s disease n=6), SLE (n=5) and healthy controls (n=12). Autoantibodies to 15,312 protein targets were measured using the GeneCopeia OmicsArray™ proteomic antigen microarray panel.

**Results:** Differential abundance analysis revealed no autoantibodies significantly elevated in GCA or other related vascular inflammatory diseases. In contrast, the SLE group showed a strong and promiscuous autoantibody response, with 175 significantly associated autoantibodies (Benjamini-Hochberg-adjusted *P* <0.05).

**Conclusions:** No autoantibodies were significantly elevated in GCA. We identified known and novel autoantibodies in SLE

**Key messages:** *What is already known on the topic?:* There is circumstantial evidence for a role of B cells in the pathogenesis of GCA. Immunohistochemical and spatial transcriptomic studies have identified B cells and plasma cells in GCA-affected arteries. Genetic studies have identified an association with the HLA class II region. Two small studies have reported autoantibodies in GCA, but these findings have not been independently validated. To date, no GCA-specific antibodies have been translated into clinical practice.

*What this study adds?:*

- Our study does not support previous reports suggesting that circulating autoantibodies are commonly observed in GCA.
- We identify known autoantibodies in SLE, providing evidence of platform validity, and in addition identify novel autoantibodies.
- We showcase a rigorous permutation-based method to evaluate whether autoantibody positivity based on thresholds derived from healthy controls in the context of high-dimensional arrays reflects the play of chance.

*How this study might affect research practice or policy?:*

- Our findings suggest that circulating autoantibodies are unlikely to be a clinically useful diagnostic in GCA.
- Further research examining roles for B cells in GCA pathogenesis independent of autoantibody production is needed.
- Future research should evaluate the newly identified SLE-associated antibodies in terms of their role in pathogenesis and their clinical utility as biomarkers of disease activity and prognosis.

## Introduction

Autoantibodies are a central feature of many rheumatic diseases and play a key role in clinical rheumatology as one of the cornerstones of diagnosis. In some immune-mediated diseases such as systemic lupus erythematosus (SLE) and Anti-Neutrophil Cytoplasmic Antibody (ANCA)-associated vasculitis, autoantibodies not only aid diagnosis but can also serve as markers of disease activity or predictors of patterns of organ involvement and prognosis^1–3^. In contrast, for giant cell arteritis (GCA), no disease-specific blood-based biomarker is currently available.

The diagnosis of GCA currently relies on a combination of clinical assessment, inflammatory markers, vascular imaging, and temporal artery biopsy. While the inflammatory markers, erythrocyte sedimentation rate (ESR) and C-reactive protein (CRP) are elevated in the majority of patients presenting with GCA, they can be raised in a wide range of inflammatory and infectious conditions and thus lack specificity. Imaging and temporal artery biopsy present logistical challenges and, importantly, their diagnostic sensitivity declines rapidly following the initiation of glucocorticoid therapy; treatment that must be commenced immediately when GCA is suspected prior to diagnostic confirmation in order to prevent irreversible sight loss^4^. Thus, there is an unmet need for more specific and accessible biomarkers in GCA.

While GCA is typically regarded as a T cell-mediated disease, several lines of circumstantial evidence have emerged that suggest that B cells may contribute to pathogenesis. The strongest genetic associations identified in GCA lie in the *HLA* class II region, implicating the presentation of self-antigens to CD4+ T cells, and, indirectly, B cells (through CD4+ T-cell help)^5^. In addition, immunohistochemical and spatial transcriptomic analyses of temporal artery tissue have demonstrated the presence of B cells, plasma cells and immunoglobulin gene expression in GCA-affected arteries^6–9^, raising the possibility that humoral immune responses may play a role in disease biology. Together, these observations led us to hypothesise that autoantibodies might be present in GCA.

Until recently, the investigation of autoantibodies has been limited by the scope of available assays, which typically measure only antibodies to either a single or a small number of predefined antigens. Of the autoantibodies which can be tested in routine clinical practice, antiphospholipid antibodies, particularly anti-cardiolipin antibodies, have been reported in approximately 30-50% of GCA patients^10–16^. However, they are not useful as a diagnostic in GCA due to lack of sensitivity or specificity. Moreover, they can occur in healthy individuals, and their prevalence rises with age in the general population^17^, which is particularly relevant to GCA since it is a disease affecting older adults. Outside of antibodies measured in routine clinical practice, two studies have reported other putative autoantibodies associated with GCA^18,19^ but small sample size and lack of independent replication mean that the robustness of these associations is uncertain. Notably, none of these reported autoantibodies have entered clinical practice despite more than a decade since publication. Advances in high-dimensional autoantibody profiling now enable unbiased screening against thousands of full-length human proteins, providing an opportunity to systematically assess the autoantibody reactome.

In this study, we applied wide-angle autoantibody profiling to patients with GCA and healthy controls, as well as inflammatory vascular diseases with similarities to GCA (Takayasu arteritis, IgG4-related disease, and Behçet’s disease with vascular manifestations). We also included systemic lupus erythematosus (SLE) to provide a disease in which autoantibodies are well-established. Using this approach, we found no evidence to support the presence of autoantibodies in GCA. In contrast, we identified a broad autoantibody repertoire in SLE, with significant increases in 175 autoantibodies compared to healthy controls, including known lupus autoantigens (e.g., Ro-52, ribosomal P, Smith, RNP) and potential novel autoantigens. Moreover, analysis at the individual patient level in SLE identified a promiscuous autoantibody response, with reactivity to over 100 autoantigen targets in each individual. Our findings do not support a role for circulating autoantibodies being commonly found in GCA and highlight the immunological distinction between GCA and classical autoantibody-mediated rheumatic diseases. We identify potential novel autoantigens in SLE that warrant further investigation in future studies.

## Methods

### Study recruitment

Serum was collected from patients with a range of immune-mediated diseases (IMDs) and healthy controls (HCs). Participants were recruited via Imperial College Healthcare NHS Trust (London, UK), Leeds Teaching Hospitals NHS Trust, the UK GCA Consortium (UKGCA) and the <u>T</u>emporal <u>A</u>rtery <u>B</u>iopsy versus <u>Ul</u>trasound in diagnosis of GCA (TABUL) study. All study participants provided written informed consent. Sample sizes were as follows: GCA (n = 20; 10 cranial GCA (C-GCA) and 10 large-vessel GCA (LV-GCA)), Takayasu arteritis (n = 12), Immunoglobulin G4-related disease (IgG4-RD, n = 5), Behçet’s disease (BD, n = 6), SLE (n = 5) and HCs (n= 12). Eight of the C-GCA patients were recruited via TABUL/UKGCA. These 8 patients were selected for this study based on high immunoglobulin (Ig) transcripts in the corresponding diagnostic temporal artery biopsy, on the basis that this might increase the likelihood of autoantibody identification. One of the C-GCA patients recruited at Imperial also had concurrent LV-GCA demonstrated on ^18^FDG-PET scan. However, systematic evaluation for large-vessel involvement was not performed and thus the true prevalence of concurrent large-vessel involvement in C-GCA patients may have been higher. Of the five of the LV-GCA patients recruited from Leeds, three underwent temporal artery biopsy (TAB) and of whom two had a positive biopsy. Five of six patients with BD had vascular manifestations (thromboses and/or arterial aneurysms) and one had a history suggestive of maternal transmission^20^. SLE samples were included to provide a disease control (since autoantibodies are a feature of this disease) and were selected to maximise autoantibody positivity to dsDNA and extractable nuclear antigens (ENA). HCs were recruited from both Imperial and Leeds and encompassed a wide span of ages to reflect the varying demographics of the different diseases in this analysis.

### Ethical approvals

For individuals recruited at Imperial College Healthcare NHS Trust, serum samples used were obtained from the Imperial College Healthcare Tissue and Biobank (ICHTB). ICHTB is supported by the National Institute for Health Research (NIHR) Biomedical Research Centre based at Imperial College Healthcare NHS Trust and Imperial College London. ICHTB is approved by Wales REC3 to release human material for research (22/WA/0214). Ethical approval was provided by the ICHTB (Human Tissue Authority Licensing number 12275). The Vasculitis and Inflammation Integrated Research Tissue Bank (VII RTB), Research Ethics Committee: Wales REC 4 (20/WA/0195), at the University of Leeds provided serum samples from Behçet’s patients, age-matched controls and TABUL (subsumed samples). TABUL was a multicentre, prospective study that compared the sensitivity and specificity of temporal artery ultrasound to biopsy in patients with suspected C-GCA (ClinicalTrials.gov identifier: NCT00974883). Additional serum samples from biopsy or imaging-confirmed GCA cases were obtained from the UK GCA Consortium (ethical approval by Yorkshire & The Humber - Leeds West Research Ethics Committee: REC 05/Q1108/28).

### Autoantibody array

Serum samples were profiled using the GeneCopeia OmicsArray™ proteomic antigen microarray panel PA014 (GeneCopeia, Rockville, Maryland, USA). This platform expresses protein antigens in *E. coli* and spotted onto nitrocellulose filter coated glass slides, providing 15,312 protein antigen targets, including 1,184 neoantigens, mapping to 6,124 unique proteins since multiple antigens were included for some proteins (**Supplementary Data 1**). The array contained technical duplicates such that each target was measured by two separate spots. IgG was purified from serum using a spin column purification using Melon-gel. 95ul of Melon buffer was used to dilute 15ul of sample, which was then loaded onto Melon-gel spin-columns and centrifuged. For the microarray analysis, fixed concentrations of purified IgG were used (0.15 mg/ml). After incubation, arrays were washed to remove any unbound antibodies. Then, to detect IgG, anti-human IgG secondary antibody (Alexa Fluor 647 goat) was added. The array was washed again to remove unbound secondary antibodies, and the slide placed into a GenePix® 4000B microarray scanner. After scanning, the GenePix® Pro 7.0 software was used to analyse raw signal data.

### Quality Control (QC) and pre-processing

After background correction and normalisation to control spots, intensity data were log_2_ transformed and quantile normalised (normalize.quantiles function, Bioconductor preprocessCore package). Spots where the duplicates had ≥ 50% coefficient of variation (CV) were removed. Duplicate spot data were then averaged. The percentage of missing values was calculated for both samples and proteins. Any samples or spots containing ≥ 50% missing values were removed from this analysis, leading to removal of 24 unique autoantigen targets. This procedure left 15,286 unique spots for analysis. Two samples (one from a patient with LV-GCA and one with BD) were removed since they were prominent outliers on principal component analysis (PCA) (**Figure S1**), suggesting they were affected by non-biological variation.

### Nomenclature

To provide a standardised nomenclature and avoid ambiguity arising from multiple protein names that refer to the same protein, each protein target was annotated using the HUGO gene symbols of the corresponding encoding gene. To retain identifiability of each specific antigen target, a unique suffix was added to each gene symbol to represent a variant of the same protein (where there was more than one antigenic target per protein).

### Differential autoantibody abundance analysis

Linear regression models (Bioconductor *limma* package) were used to compare the abundance of autoantibodies (against the 15,286 antigens passing QC) in each IMD versus healthy controls. The regression models were of the form:

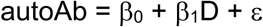

where autoAb is autoantibody abundance (continuous variable) and D is disease status (binary variable, where 0 represents HC and 1 represents the IMD being tested). β_0_ represents the intercept and β_1_ the effect size (log_2_ fold change), and ε, the residuals. To correct for multiple comparisons, Benjamini-Hochberg (BH) adjustment was applied and an adjusted p-value of < 0.05 (false discovery rate, FDR, <5%) was used to define statistical significance.

### Pathway enrichment testing of SLE associated autoantigens

We performed pathway enrichment of autoantigens corresponding to the 175 significant autoantibodies in the differential abundance analysis using overrepresentation analysis (R clusterProfiler package). Since pathway enrichment requires a single feature per protein, where there were multiple significant autoantibodies mapping to the same protein, the autoantibody with the greatest log_2_ fold change was used, resulting in 129 significant unique protein targets. These 129 proteins were tested for enrichment of Gene Ontology (GO) terms versus the background set of 6,125 protein targets measured on the array (enrichGO function, clusterProfiler package). To correct for multiple comparisons, BH adjustment was applied and an adjusted p-value of < 0.05 used to define significance.

### Defining autoantibody positivity

To binarise each autoantibody into positive and negative on a per sample basis, for each autoantigen target *j*, we defined the threshold *T_j_* for positive reactivity as > 3 standard deviations (SDs) above the mean of the HC samples for that target. We then enumerated the number of samples positive for each potential autoantibody in each IMD.

### QQplots

QQ plots were generated by plotting the observed test statistics to those expected under the null hypothesis of no association.

### Permutation analysis for empirical p-value calculation for autoantibody positivity analysis

Given the large-multiple testing burden, we used permutation testing to investigate whether the observed autoantibody positivity in a proportion of GCA patients was significant or alternatively could be explained by chance. We took the matrix of autoantibody abundance values for GCA and HCs and, keeping the numeric matrix fixed, we randomly permuted the sample labels. Using the permuted labels, new thresholds for positivity were calculated for each antigen using the same formula as described previously (now based on the samples labelled as ‘HCs’ under permutation i.e. ‘pseudo-controls’), and from this positivity of samples labelled as ‘GCA’ under permutation (‘pseudo-GCA’) was determined. For each potential autoantibody, we calculated the difference (*D_j_*) in the proportion of positives between pseudo-GCA and pseudo-controls. This permutation procedure was repeated 10,000 times. For each permutation, we defined *S* as the maximum value of *D_j_* (i.e. the maximum difference in the proportion of positivity between the pseudo-GCA group and pseudo-control group across all autoantigen targets). We then calculated an empirical p-value as follows:

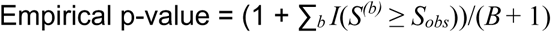

where *B* is the number of permutations performed (10,000), *S_obs_* is the maximum difference in autoantibody positivity from the real (unpermuted) data, *S(b)* is the maximum difference in autoantibody positivity for permutation *b*, and *I* is the indicator function.

The empirical p-value provides the probability of seeing a result as extreme or more extreme than the one we observed if the null hypothesis of no difference in autoantibody positivity between GCA and HC was true. It is calculated by enumerating how often we see a maximum difference in autoantibody positivity (between pseudo-GCA and pseudo-controls) in the permuted data that is greater than or equal to the maximum difference we see for GCA versus controls in the real data.

The same permutation testing approach was then performed for the SLE analysis (using the SLE and HC data).

## Results

### Study overview and clinical characteristics

To investigate whether circulating autoantibodies are present in GCA, serum samples were tested for reactivity to autoantigens using a microarray-based autoantibody profiling platform (GeneCopeia OmicsArray™ panel PA014) autoantibody microarray (**Figure 1**) and compared to healthy controls. In addition, we analysed n=5 SLE samples and samples from inflammatory vascular diseases with similarities to GCA; Takayasu arteritis (n=12), IgG4-RD (n=5), Behçet’s disease (n=6, of whom 5 had vascular involvement). Of the GCA patients, 10 had C-GCA (defined as dominant presentation with cranial symptoms and histopathologically-proven GCA on temporal artery biopsy) and 10 patients had LV-GCA (defined as dominant clinical presentation with large-vessel symptoms and confirmation of LV-GCA on ^18^FDG-PET-CT imaging). In 3 patients there was evidence of concurrent cranial and large-vessel involvement: one of the C-GCA patients had LV-GCA evident on ^18^FDG-PET-CT scan and two of the LV-GCA patients had a positive temporal artery biopsy, highlighting that C-GCA and LV-GCA are not mutually exclusive. Investigations were dictated by symptoms and thus the true prevalence of concomitant C-GCA and LV-GCA could have been higher. Demographic characteristics were typical for C-GCA and LV-GCA, with female predominance (60%) in both C-GCA and LV-GCA. C-GCA patients were older (median age 74 years) than LV-GCA (median 69 years) (**Table 1**). For GCA patients, disease duration at the time of sampling was median 0.15 years (range 0-4); for 8 of the 10 patients, samples were taken during initial presentation. As expected, patients with SLE and Takayasu arteritis were younger and had stronger female preponderance.

**Figure 1:**
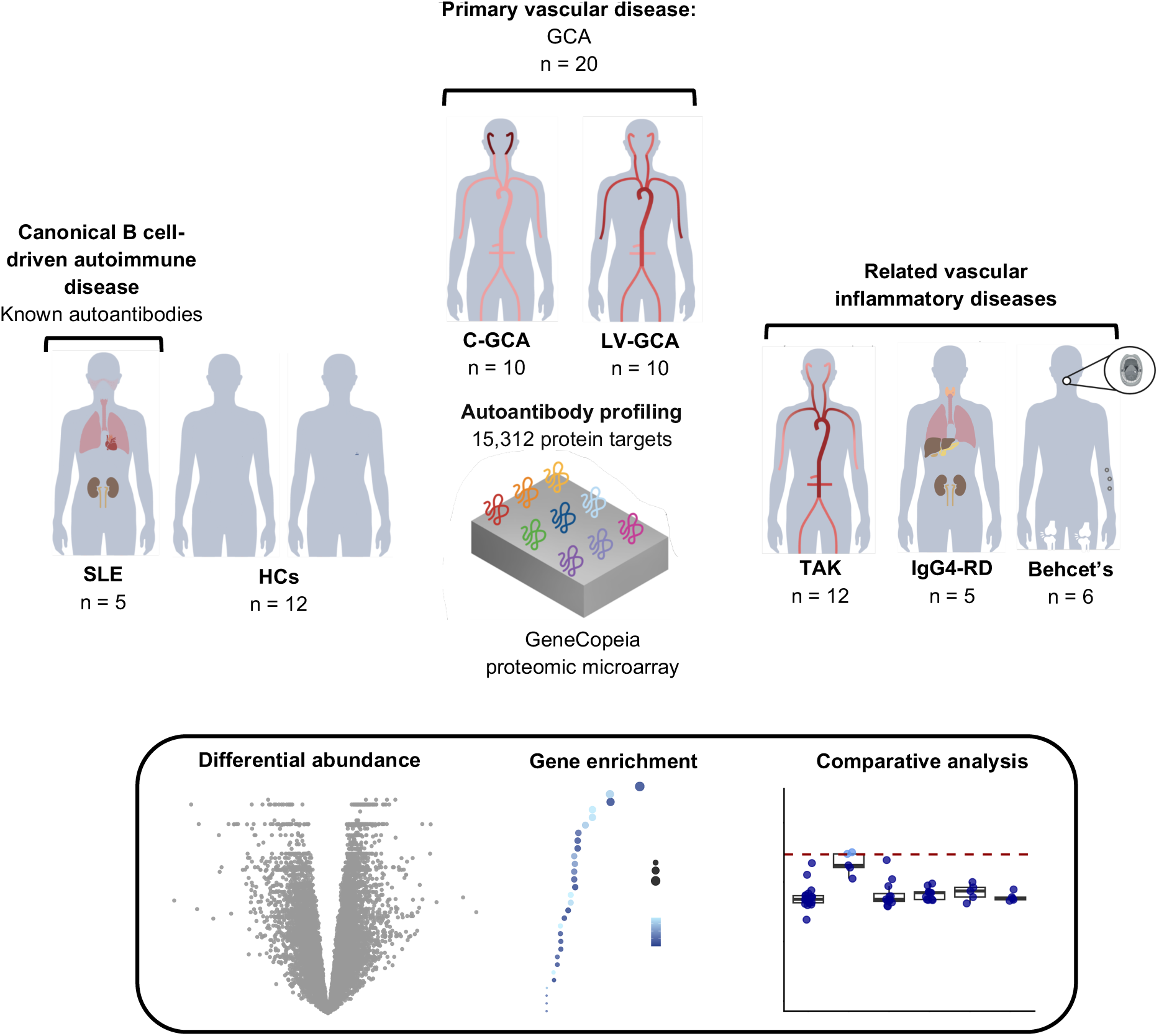
Study overview. Autoantibody profiling was performed in giant cell arteritis (GCA), healthy controls (HCs), and systemic lupus erythematosus (SLE), a disease in which the presence of autoantibodies is well-established. In addition, samples from other inflammatory vascular diseases (Takayasu arteritis (TAK), IgG4-related disease (IgG4-RD), and Behçet’s disease) were profiled. C-GCA: cranial GCA, LV-GCA: large-vessel GCA.

**Table 1:** Demographic characteristics for IMDs and healthy volunteers. * One LV-GCA sample and one BD sample were removed following QC.

| Group | Sample number (n) | N female (%) | Age (yrs) [IQR] | Disease duration (yrs) [range] |
| --- | --- | --- | --- | --- |
| GCA | 20 | 12 (60) | 72 [67, 76] | 0.15 [0-4] |
| C-GCA | 10 | 6 (60) | 74 [71, 76] | 0 [0-4] |
| LV-GCA | 10* | 6 (60) | 69 [63, 72] | 0.4 [0-3.1] |
| Healthy controls | 12 | 8 (67) | 47 [32, 77] | - |
| Takayasu arteritis | 12 | 11 (92) | 28 [26, 37] | 1.385 [0-3.3] |
| BD | 6* | 3 (50) | 42 [40, 46] | 6.5 [1-8] |
| IgG4-RD | 5 | 0 (0) | 53 [47, 68] | 0.3 [0-5.6] |
| SLE | 5 | 4 (80) | 24 [20, 27] | 0.2 [0-15.5] |

### Multi-dimensional autoantibody measurement in SLE reveals known and novel associations

B cell hyperactivity and the production of autoantibodies against nuclear components (anti-nuclear antibodies, ANA) are hallmarks of SLE pathogenesis; thus, SLE can serve as a ‘positive control’ group. Despite the small sample size of the SLE group (n=5), we observed significantly elevated reactivity to 175 autoantigen targets in SLE versus healthy controls (n=12) after adjustment for multiple testing (5% FDR) (**Figure 2a, Supplementary Data 2**). The autoantibodies with the largest log_2_ fold changes in SLE were directed against TRIM21 (Ro-52) and RPL0 (ribosomal-P), both known lupus autoantigens (**Figure 2a-b**). Among other significant autoantigens were various small nuclear ribonucleoproteins (snRNPs) including SNRPB and SNRPD3, which are targets of anti-Smith antibodies, and SNRPC, a target of anti-RNP-A autoantibodies. This provides validation of the assay technology by demonstrating that the array platform reliably detects established lupus autoantibodies. We also observed strong reactivity to SNRPN (snRNP polypeptide N), which is not a canonical target of Sm or RNP antibodies. Heatmap visualization of the 175 significant autoantibodies showed that this signal was specific for SLE and was not seen in the other vascular inflammatory diseases (**Figure S2**). Of note, one of the twelve HCs displayed a similar profile to the SLE patients, presumably representing asymptomatic antibody positivity to nuclear antigens. Pathway overrepresentation analysis of the targets of the potential 175 SLE-associated autoantibodies using Gene Ontology (GO) terms identified strong enrichment of nuclear structures and processes (e.g. ‘ribonucleoprotein complex’, ‘mRNA metabolic processes’ and ‘mRNA binding’; **Figure 2c**), consistent with the known autoreactivity to nuclear antigens in SLE and providing further validation of the platform.

**Figure 2:**
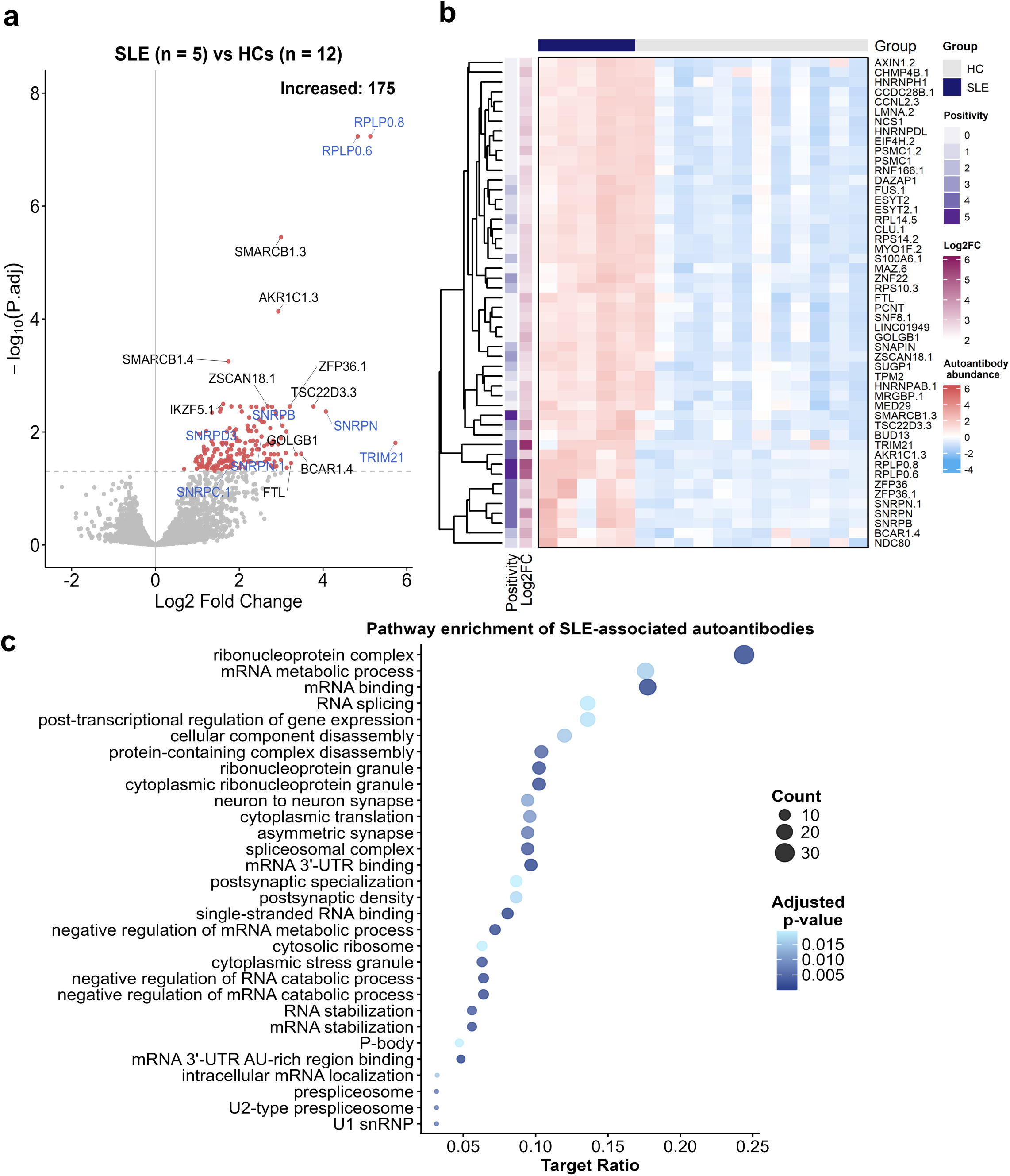
Comparison of SLE versus healthy controls. **a)** Volcano plot comparing abundance of autoantibodies against 15,286 autoantigens in SLE vs healthy controls (HCs). Each point represents a protein target on the autoantibody array. Horizontal dotted line represents significance threshold (5% FDR). Red: autoantibody significantly increased in SLE, grey: non-significant. P.adj: Benjamini-Hochberg-adjusted p-value. Selected autoantigens with the strongest log_2_ fold changes and/or −log_10_ p-values are annotated. Autoantigens are labelled by their encoding gene symbols; where multiple targets corresponding to the same protein target a period and numeric suffix are appended to the gene symbol to preserve a unique identifier. Blue gene symbols: established autoantigens in SLE measured by current clinical assays. **b)** Heatmap of the 50 autoantibodies with the greatest log_2_ fold changes. Each row (autoantibody) has been z-score normalised. Dendrogram shows hierarchical clustering of autoantibodies. ‘Positivity’: number of samples exceeding our autoantibody positivity threshold (HC mean abundance + 3 SD) for each autoantigen. ‘Log2FC’: SLE vs HC log_2_ fold change for each autoantibody. **c)** Pathway enrichment using Gene Ontology (GO) terms for the 175 significant autoantibodies. Over-representation analysis was performed against the background set of protein targets measured. Count = number of input targets (significant autoantigens) annotated to a specific pathway. Target ratio = number of significant autoantigens divided by the number of proteins in the pathway. The top 30 enriched pathways are displayed.

In addition to recapitulating clinically established lupus autoantibodies we also replicated several putative novel autoantigens reported by Lewis *et al*^21^ including FUS, HMG20B and TFE3 (**Figure S3, Supplementary Data 2**). Moreover, we also identified multiple autoantigen targets that have not been previously reported. Among the novel autoantibodies with the greatest effect sizes were those directed at including TSC22D3, BCAR1 and GOLGB1 and ZFP36. TSC22D3, also known as Glucocorticoid-Induced Leucine Zipper (GILZ), is a potent anti-inflammatory molecule whose expression is induced by glucocorticoids and appears to be protective in SLE^22^. Like GILZ, ZFP36 is a negative regulator of inflammation. ZFP36 is an RNA-binding protein that promotes the degradation of mRNA of pro-inflammatory cytokines including TNF and IL-6^23,24^

The differential autoantibody abundance analysis tests for differences in group means between SLE and healthy controls. Although this analysis framework is the most statistically rigorous approach, we also wished to evaluate how autoantibodies varied at the individual level and to explore potential for clinical translation by defining an autoantibody as positive or negative (as currently done in clinical practice). Autoantibodies where the distribution of values are minimally overlapping with HCs will be most promising for translational potential in terms of discriminating SLE from HCs. Therefore, for each autoantibody, we defined positivity as a value > 3 SDs above the mean of the HC samples. We then enumerated the proportion of SLE samples positive for each antibody. Using this definition, we observed that the number of autoantibodies in any given patient with SLE was greater than 100, and in one patient more than 500 autoantibodies were detected (**Figure 3a**). Of the clinically established SLE-associated autoantibodies, anti-ribosomal P were positive in all 5 SLE samples, anti-Ro-52 in 4 samples and anti-SNRPB in 4 samples (**Figure 3b-e**). Cross-referencing with ENA profiles measured in the clinical laboratory revealed high concordance between positivity on the microarray and the reference values (**Figure S4**). Among the novel autoantibodies with the strongest log_2_ fold changes in the differential abundance analysis, SMARCB1 (ranked 16^th^ by log_2_ fold change) was positive in all 5 samples, ZFP36 in 4 samples and TSC22D3 (GILZ) in 3 samples (**Figure 3g-h**). Notably, for some autoantibodies (e.g. GOLGB1 and FTL) identified during the differential autoantibody analysis, no SLE samples were positive based on the threshold we implemented here, despite high ranking by log_2_ fold changes in the primary analysis. Visualisation of these examples confirmed that while abundance of these antibodies was indeed higher in SLE than HCs, no sample met our positivity threshold of >3 SD from the mean of HCs (**Figure S5**).

**Figure 3:**
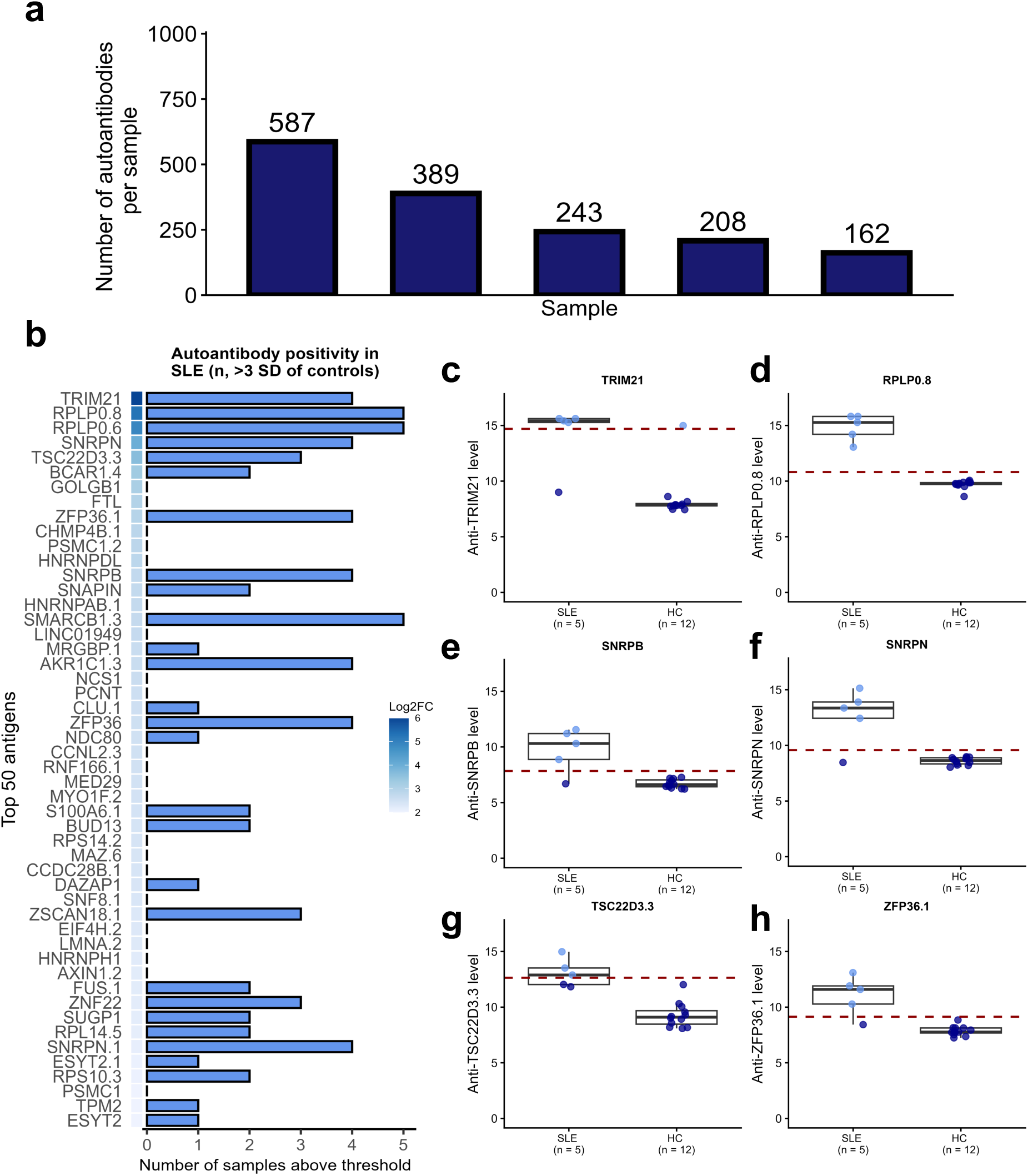
The autoantibody reactome in SLE. **a)** The number of autoantibodies exceeding the positivity threshold (HC mean + 3 SD) per sample. **b)** Number of SLE samples exceeding the autoantibody positivity threshold for the top 50 autoantigens ranked by SLE vs HC log_2_ fold change (vertical bar ‘Log2FC’). **c-h)** Box-and-whisker plot showing examples of specific clinically established **(c-e)** and novel **(f-h)** autoantibodies in SLE. Box lower and upper bounds represent first and third quartile. Middle bar within box = median. The red horizontal dotted line represents the autoantibody positivity threshold. Light blue and dark blue points = above and below positivity threshold, respectively.

### No evidence for circulating autoantibodies in GCA

We next addressed our main research question, testing for autoantibodies in GCA. Differential autoantibody abundance analysis comparing GCA (n = 19) versus HCs (n = 12) showed no significant differences (**Figure 4a, Supplementary Data 3**). Given that the sample size of the GCA was approximately four-fold that of SLE (in which we identified 175 significant associations), it seemed unlikely that lack of statistical power could account for lack of significant associations for common autoantibodies. Nevertheless, to explore this further we used QQplots to examine the distribution of test statistics. The observed p-values from the GCA versus HC comparison closely followed the expected uniform distribution under the null hypothesis, providing no evidence to support an autoantibody signal in GCA (**Figure 4b**). In contrast, the observed p-values obtained from SLE versus HC deviated from the expected distribution under the null hypothesis (**Figure 4b**).

**Figure 4:**
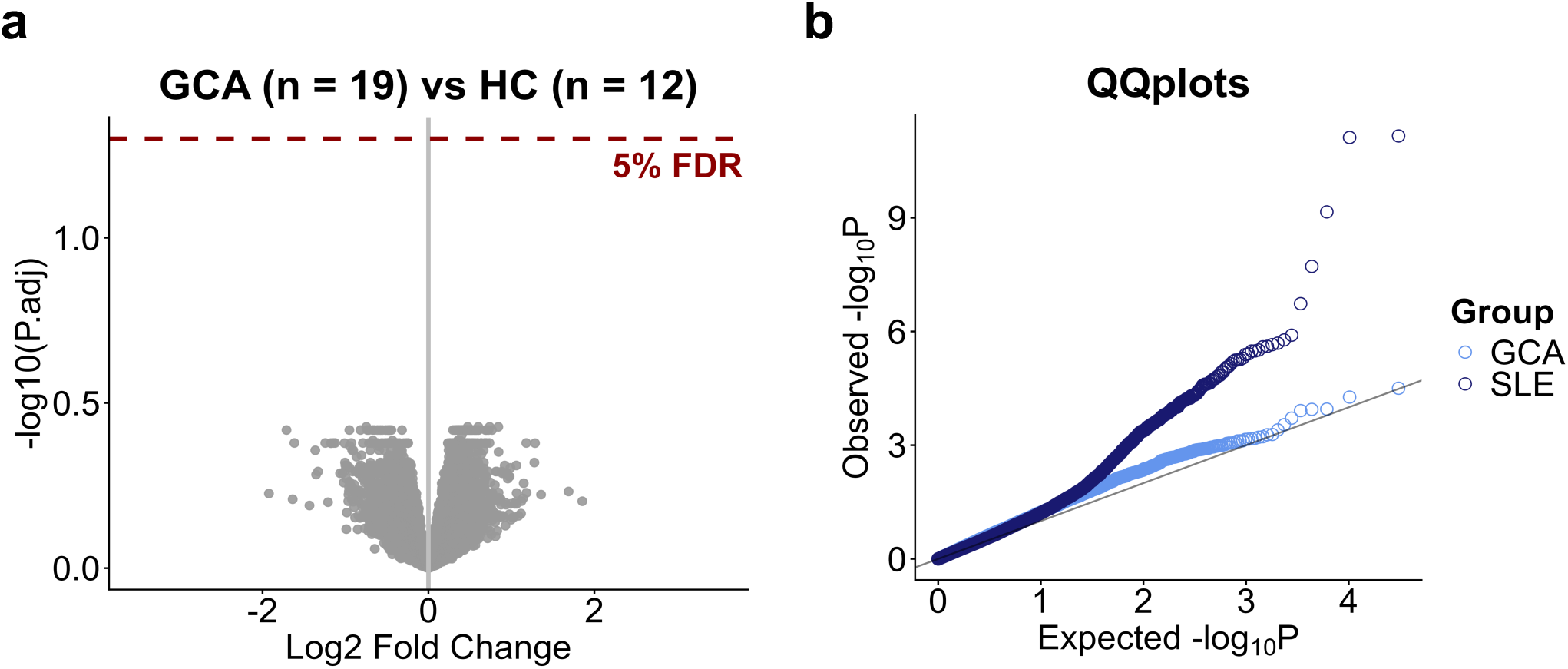
No evidence of autoantibodies in GCA. **a)** Volcano plot comparing abundance of autoantibodies against 15,286 autoantigens in GCA versus HCs. Each point represents a protein target. Horizontal dotted line represents 5% FDR significance threshold**. b)** QQplots examining the distribution of observed test statistics against those expected under the null hypothesis. Light blue: GCA versus HC, dark blue: SLE versus HC. Diagonal line represents line of identity.

Some investigators have reported differences in the presence of B cell infiltrates in aortic tissue from LV-GCA versus temporal arteries from C-GCA^6^. We therefore considered the possibility that, hypothetically, there might be differences in autoantibodies in the LV-GCA versus C-GCA, and that, if so, analysing them as a single entity could mask subgroup-specific autoantibodies. We therefore repeated differential autoantibody abundance analysis separately for LV-GCA (n=9, since 1 sample was removed during QC) and for C-GCA (n=10) versus HCs (n=12). In line with the analysis of all GCA versus HCs, no significant differences were observed for either the LV-GCA or the C-GCA subgroup analyses (**Figure S6, Supplementary Data 4-5**).

There have been a small number of previous studies that have reported the identification of autoantibodies in GCA^15,16,18,19,25^. Six of these protein targets were included on the GeneCopeia microarray used in the present study (some of these proteins were represented by multiple isoforms). These were: Lamin A/C (LMNA), Vinculin (VCL), Voltage Dependent

Anion Channel 2 (VDAC2), Far Upstream Element-Binding Protein 2 (FUBP2; also known as KH-Type Splicing Regulatory Protein (KHSRP)), Growth Factor Receptor Bound Protein 2 (GRB2), Ferritin Heavy Chain 1 (FTH1). Though cardiolipin could not be examined on this array since it is a phospholipid rather than a protein, β2-glycoprotein I (β₂GPI, also known as apolipoprotein H (APOH)) was included on the array. No antibodies to any of these targets were statistically significant in our analysis. Moreover, none had even a nominal P-value <0.05 (i.e. uncorrected P-value prior to multiple testing adjustment). We then enumerated the number of GCA samples with positivity to these antibodies using the definition described earlier. Autoantibody positivity was restricted to no more than two samples and in most cases was absent (**Table 2**). In summary, our data do not replicate previous studies identifying these autoantibodies in GCA.

**Table 2:** Evaluation of autoantibodies reported in previous studies. Autoantibody positivity threshold defined as mean abundance + 3 SD of the healthy control group. Nominal p-value: P from limma before multiple testing adjustment. BH=Benjamini-Hochberg. LMNA=Lamin A/C, VCL=Vinculin, VDAC2=Voltage Dependent Anion Channel 2, FUBP2=Far Upstream Element-Binding Protein 2 (also known as KH-Type Splicing Regulatory Protein (KHSRP)), GRB2=Growth Factor Receptor Bound Protein 2, FTH1=Ferritin Heavy Chain 1, APOH = Apolipoprotein H (also known as β_2_-glycoprotein 1).

| Antigen | HGNC.ID | Study | PudMed ID | Variant | Number GCA samples autoantibody positive (%) | Differential autoantibody abundance testing GCA vs HCs |  |
| --- | --- | --- | --- | --- | --- | --- | --- |
|  |  |  |  |  |  | Nominal p-value | BH-adjusted p-value |
| LMNA | 6636 | Régent (2011) et al<sup>19</sup> | 21711540 |  |  |  |  |
|  |  |  |  | LMNA | 0 (0%) | 0.64 | 0.90 |
|  |  |  |  | LMNA.1 | 0 (0%) | 0.54 | 0.88 |
|  |  |  |  | LMNA.2 | 0 (0%) | 0.76 | 0.95 |
|  |  |  |  | LMNA.3 | 2 (10.5%) | 0.99 | 1.00 |
|  |  |  |  | LMNA.4 | 0 (0%) | 0.75 | 0.95 |
|  |  |  |  | LMNA.5 | 0 (0%) | 0.22 | 0.73 |
|  |  |  |  | LMNA.6 | 1 (5.3%) | 0.58 | 0.89 |
|  |  |  |  | LMNA.7 | 1 (5.3%) | 0.42 | 0.83 |
| VCL | 12665 |  |  |  |  |  |  |
|  |  |  |  | VCL | 2 (10.5%) | 0.13 | 0.68 |
|  |  |  |  | VCL.1 | 0 (0%) | 0.34 | 0.80 |
|  |  |  |  | VCL.2 | 0 (0%) | 0.56 | 0.89 |
| VDAC2 | 12672 |  |  |  |  |  |  |
|  |  |  |  | VDAC2 | 0 (0%) | 0.38 | 0.81 |
| FUBP2 | 6316 |  |  |  |  |  |  |
|  |  |  |  | KHSRP | 1 (5.3%) | 0.58 | 0.89 |
|  |  |  |  | KHSRP.1 | 0 (0%) | 0.81 | 0.96 |
|  |  |  |  | KHSRP.2 | 1 (5.3%) | 0.16 | 0.70 |
|  |  |  |  | KHSRP.3 | 0 (0%) | 0.054 | 0.58 |
| GRB2 | 4566 |  |  |  |  |  |  |
|  |  |  |  | GRB2 | 2 (10.5%) | 0.15 | 0.70 |
|  |  |  |  | GRB2.1 | 0 (5.3%) | 0.66 | 0.91 |
| FTH1 | 3976 | Baerlecken (2012) et al<sup>18</sup> | 22228484 |  |  |  |  |
|  |  |  |  | FTH1 | 0 (0%) | 0.35 | 0.80 |
|  |  |  |  | FTH1.1 | 0 (0%) | 0.78 | 0.95 |
|  |  |  |  | FTH1.2 | 0 (0%) | 0.92 | 0.99 |
|  |  |  |  | FTH1.3 | 0 (0%) | 0.54 | 0.88 |
|  |  |  |  | FTH1.4 | 0 (0%) | 0.007 | 0.42 |
|  |  |  |  | FTH1.5 | 0 (0%) | 0.84 | 0.97 |
|  |  |  |  | FTH1.6 | 0 (0%) | 0.20 | 0.72 |
|  |  |  |  | FTH1.7 | 0 (0%) | 0.38 | 0.81 |
|  |  |  |  | FTH1.8 | 0 (0%) | 0.16 | 0.70 |
| APOH | 616 | Espinosa (2001) et al<sup>16</sup> |  |  |  |  |  |
|  |  |  |  | APOH | 0 (0%) | 0.06 | 0.60 |
|  |  |  |  | APOH.1 | 0 (0%) | 0.25 | 0.75 |

Takayasu arteritis is a rarer form of large-vessel vasculitis (LVV). It has similarities to GCA in terms of histopathology (granulomatous arteritis), arteries involved (e.g. aorta and its branches) and circulating inflammatory protein profile^26^, although there are also distinctions in terms of patient demographics, genetic aetiology and arterial tropism. Differential autoantibody abundance testing of Takayasu arteritis (n=12) versus HCs (n=12) revealed no associations (**Figure S7, Supplementary Data 6**). Analysis of a combined LVV group (GCA and Takayasu arteritis) versus HCs again revealed no significant associations (**Supplementary Data 7**).

We had also collected samples from other inflammatory vascular diseases (BD and IgG4-RD; the purpose of this had been that in the event of identifying GCA-associated autoantibodies, we wished to test whether these were specific to GCA or non-specific markers of vascular inflammation and/or injury). Differential autoantibody testing of BD versus HCs and of IgG4-RD versus HCs did not reveal any significant associations (**Figure S7, Supplementary Data 8-9**). However, power was limited and our intent in profiling these samples had not been a primary discovery analysis and thus this result should be interpreted cautiously.

### Autoantibody positivity in GCA likely reflects play of chance

We systematically evaluated antibody positivity for GCA samples across all measured autoantigens, using the same approach based on the distribution of the HC samples as described earlier. Using this definition, for any given autoantibody, the majority of patients with GCA were negative. Where we did observe positive autoantigen reactivity, in the large majority of instances just a single patient was positive against that target (**Figure S8**). The maximum prevalence of autoantibody positivity in GCA was 36.8% (7 of 19 patients), which occurred for four autoantigens (CCDC47, PDE4DIP, RAF1, ZFP36). In contrast, in SLE there were 3 autoantibodies that were positive across all patients. The comparison of these distributions is shown in **Figure S8**, although a caveat is the small SLE sample size (n=5). Of note, the ZFP36 antigen target (amino acids 236-326) exhibiting reactivity about the positivity threshold in these 7 GCA samples was distinct to the ZFP36 antigenic target in the SLE analysis (amino acids 1-176, containing zinc-finger domains) described earlier.

Unlike for the differential abundance analysis, where we applied the Benjamini-Hochberg correction to control the false discovery rate at 5%, the analysis of autoantibody positivity on a per sample basis described above does not account for multiple testing. Given the high multiple testing burden (over 15,000 tests), we reasoned that autoantibody positivity based on a definition of greater than the mean + 3 SD of the HCs for that autoantibody might simply reflect play of chance. To formally evaluate this, we used a permutation procedure keeping the numeric matrix of autoantigen reactivity unchanged but randomly shuffling the sample labels each permutation and repeating this over 10,000 permutations. The permutation procedure breaks up any potential relationship between phenotype and the molecular data, enabling empirical estimation of the null distribution without any a priori assumptions. For each permutation, the maximum difference in positivity between the pseudo-GCA group (i.e. the group labelled as GCA under permutation of sample labels) and the pseudo-control group was calculated, enabling generation of the distribution of test statistics under the null hypothesis of no difference in autoantibody positivity rate between GCA and HCs. An empirical p-value was then calculated by comparing the observed test statistic (maximum difference in positivity between disease and HCs in the real data) with its null distribution. For GCA, the empirical p-value was non-significant (P 0.84), indicating that the observed patterns in autoantibody positivity in GCA would be expected under the null hypothesis. In contrast, for the SLE group, the empirical p-value was significant (P 0.043), indicating that the observed patterns in autoantibody positivity are unlikely to have arisen by chance. These findings align with those of our differential abundance autoantibody abundance analysis using linear models. While the two approaches test different hypotheses, they lead to the same conclusion: in this dataset, there is strong evidence for multiple autoantibodies in SLE and no evidence for autoantibodies in GCA.

## Discussion

GCA has traditionally been considered a T cell-mediated disease, and historical immunohistochemical studies reported few, if any, B cells in GCA-affected temporal arteries^6,7,27,28^. More recently several lines of circumstantial evidence have emerged that suggest that B cells may contribute to GCA pathogenesis. B cells and plasma cells have been reported in temporal artery biopsy and aortic samples from patients with GCA, particularly in the adventia^6,7,29^. In LV-GCA aortic tissue, B cell infiltrates can form organised arterial tertiary lymphoid organs (ATLOs) containing germinal centers, proliferating B cells and localised IgG+ plasma cell niches^6^, while a spatial transcriptomic study of GCA TABs reported adventitial enrichment of IgG-producing plasma cells^8^.

In the present study, we used a high-dimensional protein microarray to interrogate the autoantibody repertoire in GCA and found no statistically significant differences in autoantibody reactivity between patients with GCA and healthy controls. While previous work has suggested that B cells may play a more prominent role in aortic pathology in LV-GCA compared with C-GCA^6^, our stratified analyses failed to identify any autoantibody associations in either disease subgroup, consistent with our primary analysis. While the absence of detectable associations does not definitively exclude the presence of disease-relevant autoantibodies, the distribution of test statistics observed in QQ plots argues against a latent autoantibody signal in our dataset. Collectively, these findings provide no support for circulating autoantibodies in the majority of GCA patients.

Notably, we were unable to replicate previously reported autoantibodies in GCA, including anti-ferritin antibodies. Anti-ferritin antibodies have been previously identified in GCA by two groups^18,30^. Baerlecken et al used a high-density proteomic microarray followed by single-plex enzyme-linked immunosorbent assay (ELISA) and detected antibodies against the human ferritin heavy chain (N-terminus) in 33 of 36 patients with untreated GCA and/or polymyalgia rheumatica^18^. However, the presence of anti-ferritin antibodies in GCA was non-specific and also detected in SLE (in 11 of 38 patients), Churg-Strauss syndrome (7 of 21 patients), and Microscopic Polyangiitis (6 of 21 patients). Regent et al also examined antibodies to the same N-terminal ferritin heavy chain peptide^30^. They observed anti-FTH1 antibodies in 72.5% of (29 of 40) patients with biopsy-confirmed GCA and 41.3% of biopsy-negative GCA patients. This study also identified anti-ferritin antibodies in 32% of patients who presented with symptoms suggestive of possible GCA, but who were ultimately diagnosed with another condition (13 of 38 patients)^30^, indicating that anti-ferritin antibodies are unhelpful as a diagnostic. While the microarray we used in the present study did not contain the precise isolated N-terminal peptide (19-45) used in these analyses, it contained 9 different epitopes for the FTH1 antigen including the full-length ferritin heavy chain. In our data, no GCA samples exceeded the positivity threshold for any ferritin antibodies and there was no significant elevation in autoantibody abundance compared to HCs (even at nominal P <0.05, i.e. before multiple testing adjustment). Of note Regent et al used a more liberal definition of positivity (mean of HCs + 2 SDs) than in our study (mean of HC + 3 SD), which may account for the high levels of anti-FTH1 antibodies seen in their study in both GCA and non-GCA samples.

Another study utilising a combination of immunoblotting and mass spectrometry has reported autoantibodies against endothelial and vascular smooth muscle cell antigens in the sera of three patients with GCA^19^. Some antibody targets found in GCA sera, and not HCs included, LMNA, VCL, FUBP2, and several mitochondrial antigens (e.g., VDAC2). In our data, very few samples showed autoantibody positivity to these antigens (maximum 2 of 19 samples for certain epitopes) and we found no statistically significant elevation in autoantibody abundance compared to HCs (even before multiple testing adjustment). Although we cannot exclude the possibility of methodological differences (such as epitope presentation, post-translational modifications, or assay sensitivity) accounting for inter-study discrepancies, the lack of replication in our analysis suggests that potential false-positive associations and overly liberal thresholds for defining antibody positivity may underlie some earlier reports.

Anti-phospholipid, particularly anti-cardiolipin, autoantibodies have been reported in GCA in several studies^10–16,25,31^. Using a single-plex ELISA, Espinoza et al revealed significantly more anti-cardiolipin antibodies in the sera of GCA/PMR patients (16 of 20) compared to HCs (10 of 50)^14^. However, these antibodies lack specificity; they are most strongly associated with primary antiphospholipid antibody syndrome and with SLE but can also be detected in other autoimmune rheumatic diseases^32–34^. In addition, they may be transiently positive after infection^35^. Their incidence in otherwise healthy individuals also rises with age^17^. In GCA their presence may reflect an incidental age-related finding or an epiphenomenon secondary to arterial injury rather than an underlying pathogenic process. Consistent with this, pathological anti-phospholipid antibodies are associated with venous and arterial thrombosis, which are not typical features of GCA. In our study, anti-cardiolipin antibodies were not measured since cardiolipin is a phospholipid and therefore was not present on this protein array. However, no GCA samples exceeded the autoantibody positivity threshold for β2GP1, a protein target of anti-phospholipid antibodies. Our finding is consistent with the study of Liozon et al, who found that anti-β2GP1 antibodies were undetectable in 45 GCA patients^12^.

We also found no evidence for disease-associated autoantibodies in Takayasu arteritis, another form of large-vessel vasculitis with overlapping clinical and pathological features. There have been previous reports of autoantibodies in Takayasu arteritis including anti-endothelial cell antibodies (AECA), anti-endothelial protein C-receptor (EPCR) and scavenger receptor class B type 1 (SR-BI)^36–38^, although these have not been robustly validated in independent cohorts and do not have a sufficiently high prevalence to make them clinically useful. Combining patients with GCA and Takayasu arteritis into a unified large-vessel vasculitis cohort did not reveal any significant autoantibody associations. Together, our data argue against a major role for circulating autoantibodies in the pathogenesis of large-vessel vasculitis.

While autoantibody production is an important aspect of B cell driven immune-mediated disease pathogenesis, B cells could potentially contribute to pathogenesis through other mechanisms including antigen presentation and cytokine production. For example, B cells from GCA-affected arteries express pro-inflammatory cytokines, and *in vitro* experiments involving stimulation of macrophages with supernatants from cultured B cells skew macrophage polarisation towards a pro-inflammatory phenotype^7^. Thus, our findings do not preclude the possibility of a role for B cells in GCA. On the other hand, the presence of B cells, plasma cells and immunoglobulin transcripts in GCA-affected arteries does not necessarily imply a pathogenic role.

We included SLE to provide a disease in which autoantibodies are known to occur. Despite the small sample size of the SLE group (n=5), we observed significantly elevated reactivity to 175 autoantigens. None of these 175 autoantibodies were significantly elevated in the other systemic inflammatory diseases analysed here. This suggests their specificity for SLE, with the caveat that our study did not include the autoimmune diseases most closely related to SLE such as Sjogren’s syndrome and systemic sclerosis. Among the strongest signals in SLE were antibodies to TRIM21 (Ro-52), RPL0 (ribosomal-P) and various small nuclear riboproteins (SNRPs) which include the targets of anti-Smith and anti-RNP antibodies. These expected ‘positive control’ hits provide important validation of the assay technology by demonstrating that it reliably detects known lupus autoantibodies. Pathway overrepresentation analysis identified strong enrichment of nuclear structures and processes, further validating the platform.

A striking finding was that the number of autoantibodies in any given patient was greater than 100, and in one patient more than 500 autoantibodies were detected. These observations suggest that the lupus autoantibody reactome is more extensive than currently recognised. However, we highlight that the SLE patients in this study were selected to maximise positivity to multiple ENAs and so further investigation is needed to establish the generalisability of this finding.

Our data suggest several novel autoantibody associations. One intriguing example was anti-TSC22D3 antibodies. TSC22D3, also known as Glucocorticoid-Induced Leucine Zipper (GILZ), is a potent anti-inflammatory molecule whose expression is induced by glucocorticoids^39^. Reduced *GILZ* mRNA and protein expression, and an impaired GILZ response to glucocorticoid treatment, has been linked to increased disease activity and higher interferon stimulated gene (ISG) expression score in patients with SLE^40^. Similarly, GILZ deletion in lupus mouse models results in exacerbated disease^22^. These data suggest that GILZ is a protective host factor in SLE. As a result, GILZ analogues are currently under development as safer glucocorticoid-like therapies with fewer metabolic toxicities, underscoring the translational relevance of this pathway. Another example of a novel autoantigen was ZFP36. Like GILZ, ZFP36 is a negative regulator of inflammation. ZFP36 is an RNA-binding protein that promotes the degradation of mRNA of pro-inflammatory cytokines including TNF and IL6^23,24^. Therefore, one could hypothesise that anti-ZFP36 antibodies might be pro-inflammatory. Thus, our preliminary data demonstrating the presence of anti-GILZ and anti-ZFP36 autoantibodies in SLE patients raise the possibility that such antibodies may neutralise endogenous ‘brakes’ on inflammation and could contribute to more severe or treatment-refractory disease.

There are some limitations of our study. It is possible that there were autoantibodies that were below level of detection on the platform used here. For example, in minimal change disease, anti-nephrin antibodies are not always reliably detectable in serum by conventional ELISA leading to false negatives when compared to immunoprecipitation^41,42^. The bacterial expression system used here expresses proteins without post-translational modifications (PTMs). PTMs such as citrullination and glycosylation can affect immunogenicity (for example, anti-citrullinated protein antibodies in rheumatoid arthritis and antibodies to the glycosylated hinge region of IgA in IgA nephropathy)^43,44^. We cannot exclude the possibility that we did not identify weak autoantibody signals due to incomplete power. However, we note that we identified strong autoantibody associations in SLE despite approximately one quarter of the sample size of GCA. Moreover, QQ plots did not suggest a latent signal in the GCA group. While the protein microarray used here contained over 15,000 protein antigen targets mapping to over 6,000 proteins, not all possible antigens/epitopes were included and thus we cannot rule out the possibility of unidentified autoantibodies in GCA. While our data in SLE demonstrates established and novel associations, we did not design this study for autoantibody discovery in SLE. A future study involving larger sample size and replication of findings with another assay type (e.g. ELISA) is required to demonstrate whether the potential novel SLE autoantibody associations we have identified are robust, and to examine associations of these antibodies with disease phenotype and prognosis.

A strength of our study was the use of a permutation approach to rigorously address whether autoantibody positivity based on healthy control-derived thresholds was simply the play of chance. As high-dimensional autoantibody arrays become more frequently used there is a pressing need for the field to adopt robust statistical approaches to avoid false positives arising from multiple testing. Definitions of antibody positivity based on the distribution of healthy control samples need to account for this issue.

In summary, we find no evidence of commonly occurring autoantibodies in GCA. We identify both known and novel autoantibodies in SLE.

## Data Availability

All data produced in the present study are available upon reasonable request to the authors.

## Acknowledgements

We gratefully acknowledge our dear colleague the late Prof. Justin Mason, who sadly passed away in 2022, for contributions to patient recruitment. We thank Prof. Marina Botto for providing samples and for comments on the manuscript.

## Supplementary Figures

**Figure S1:**
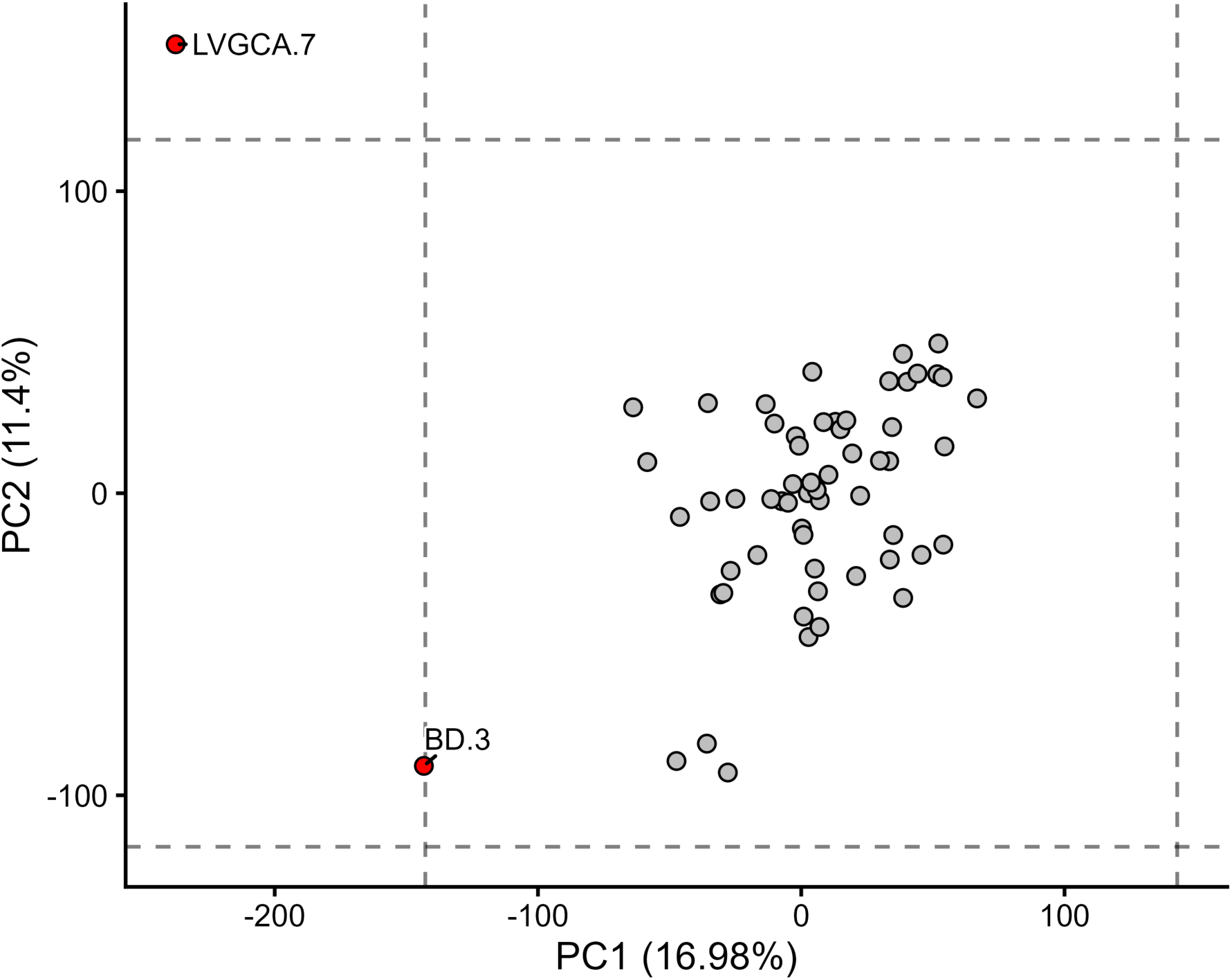
Outlier detection using principal component analysis (PCA). Each point represents a sample. Vertical dashed line = mean of PC1 values ± 3 standard deviations (SDs). Horizontal dashed line = mean of PC2 ± 3 SDs. Points (red) outside the bounds of the resulting box were considered outliers.

**Figure S2:**
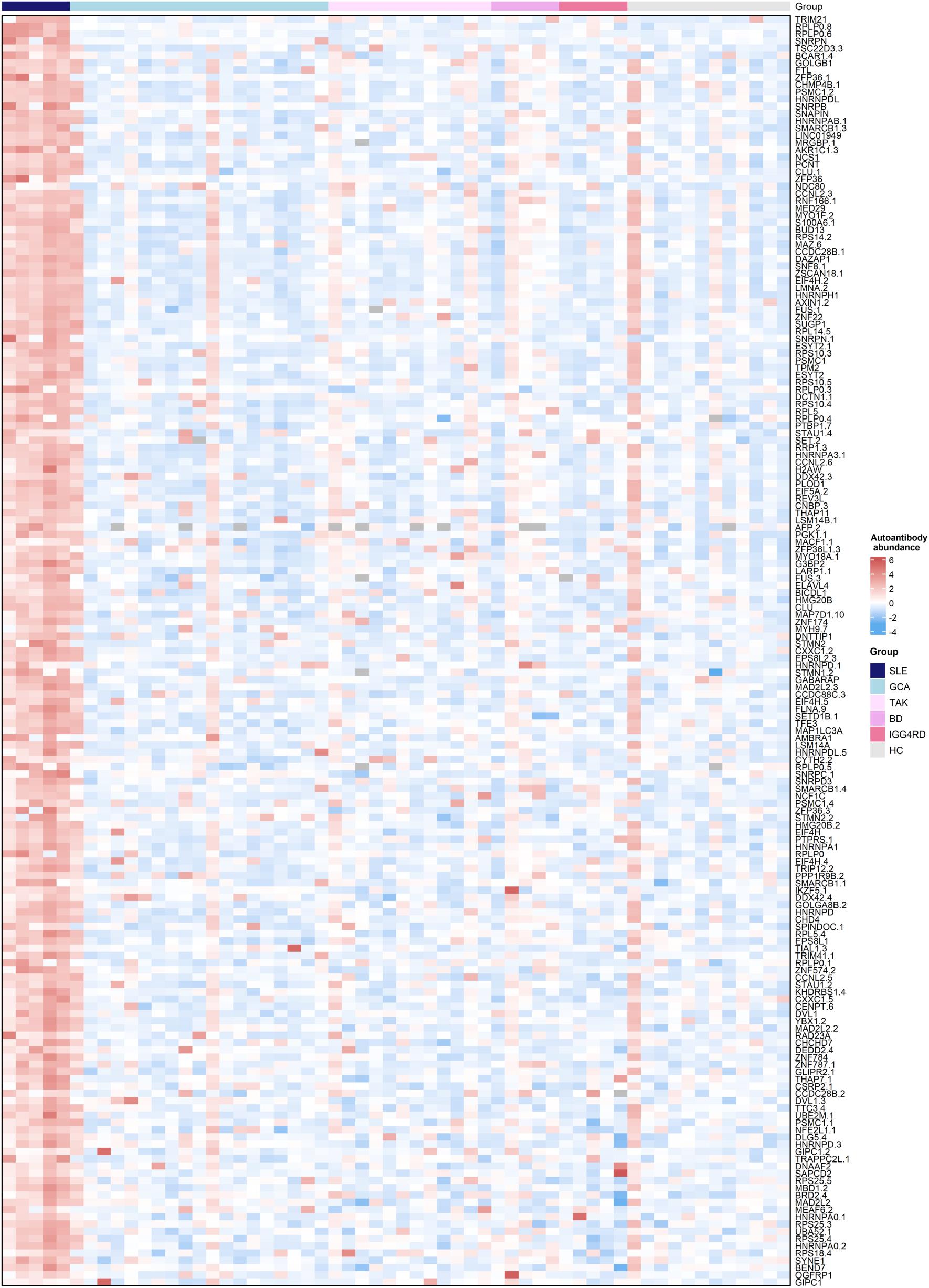
Heatmap of the 175 autoantibodies significant in the SLE vs HC differential abundance analysis. Autoantibodies (rows) have been ordered from greatest to lowest log_2_ fold change. Each row has been z-score normalised. Horizontal upper bar denotes disease status.

**Figure S3:**
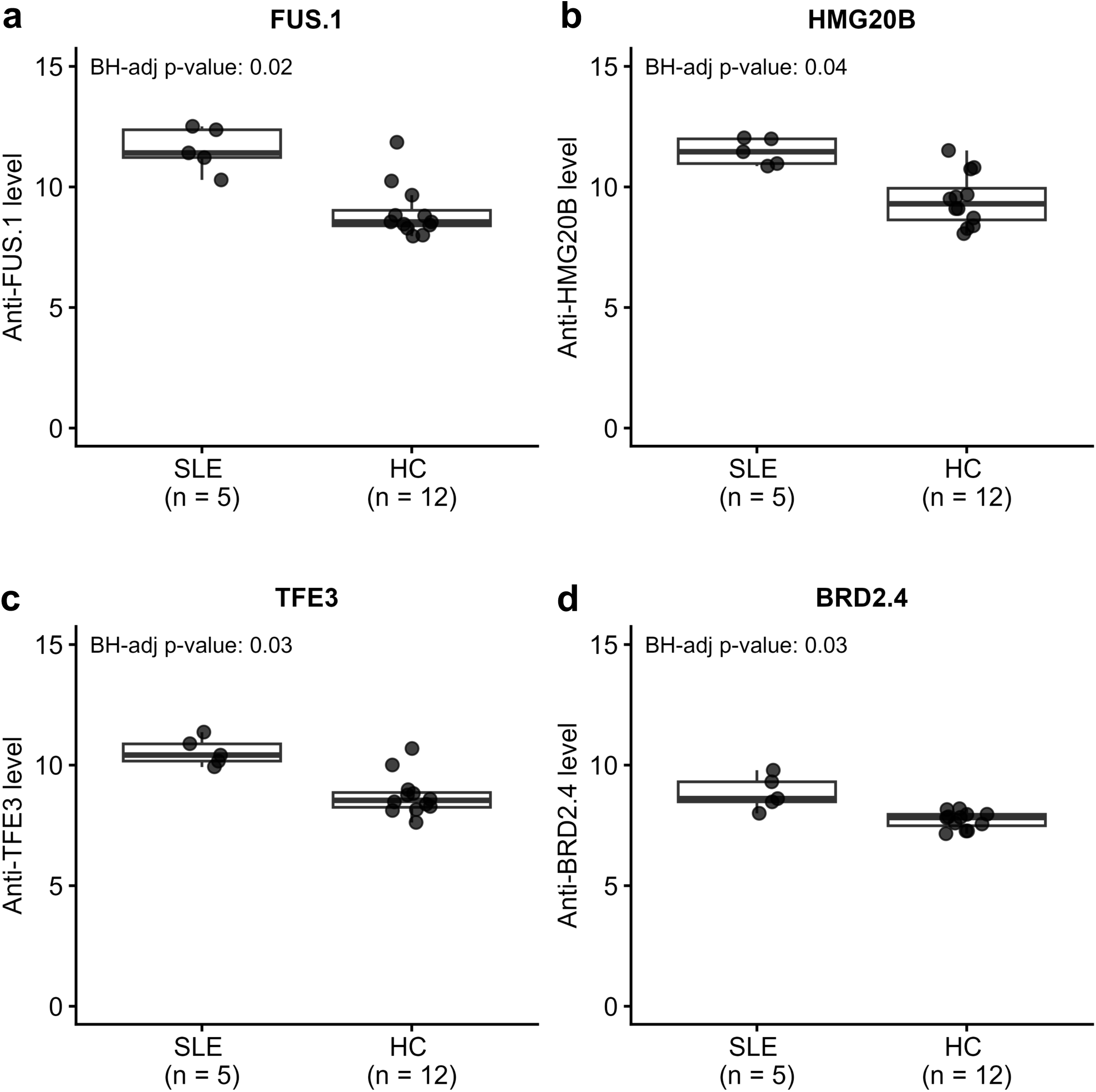
Replication of SLE-associated autoantibodies from Lewis et al^21^. **a-d)** Box-and-whisker plots showing autoantibodies with significant differential abundance in the SLE vs HC comparison in the present study and Lewis et al^21^. Box lower and upper bounds represent first and third quartile. Middle bar within box = median. BH-adj p-value= Benjamini-Hochberg adjusted p-value.

**Figure S4:**
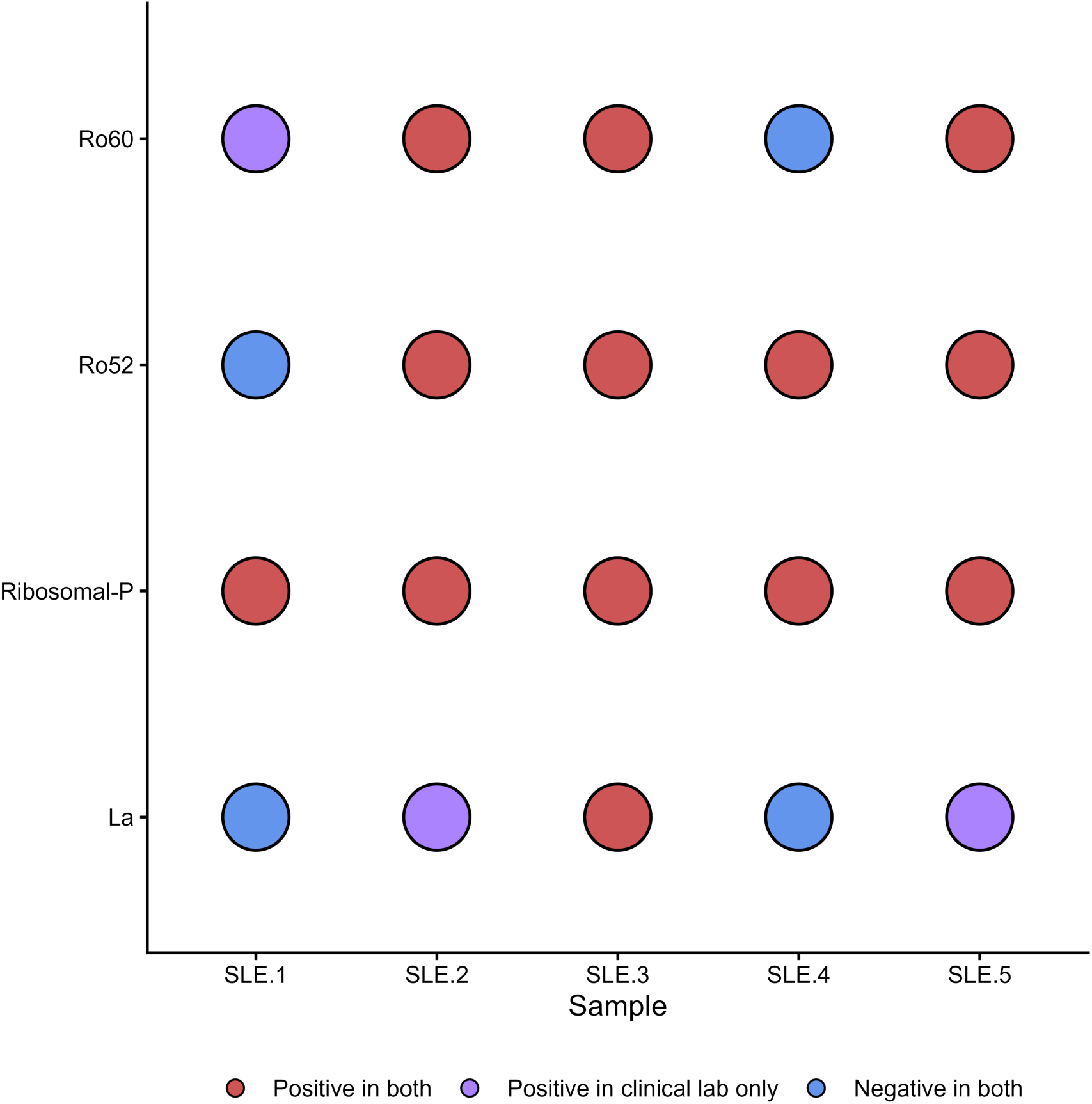
Concordance of clinical lab measurements vs the GeneCopeia microarray for autoantibodies to Extractable Nuclear Antigens (ENA). For each SLE sample, concordance of clinical laboratory and microarray-measured autoantibody status is shown. Data for Sm and RNP antibodies are not shown here because the corresponding antigens are multi-protein complexes which map to numerous targets on the microarray complicating visual presentation.

**Figure S5:**
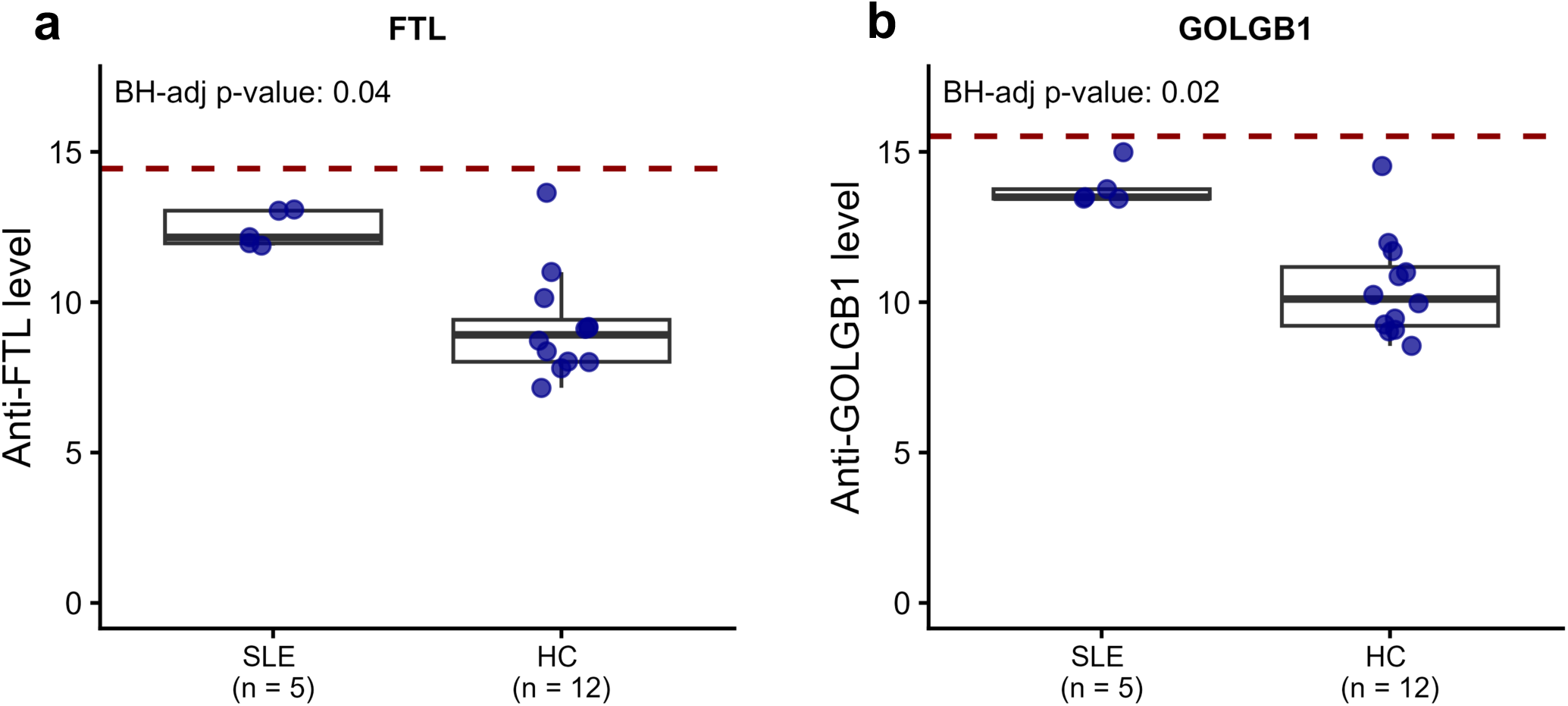
Significant elevation in group mean of SLE vs HCs does not always translate into autoantibody positivity. **a-b)** Box-and-whisker plot showing autoantibodies that were significant in the SLE vs HC differential abundance analysis, but where no SLE sample met our defined threshold for autoantibody ‘positivity’. The red horizontal dashed line represents the autoantibody positivity threshold. Box lower and upper bounds represent first and third quartile. Middle bar within box = median.

**Figure S6:**
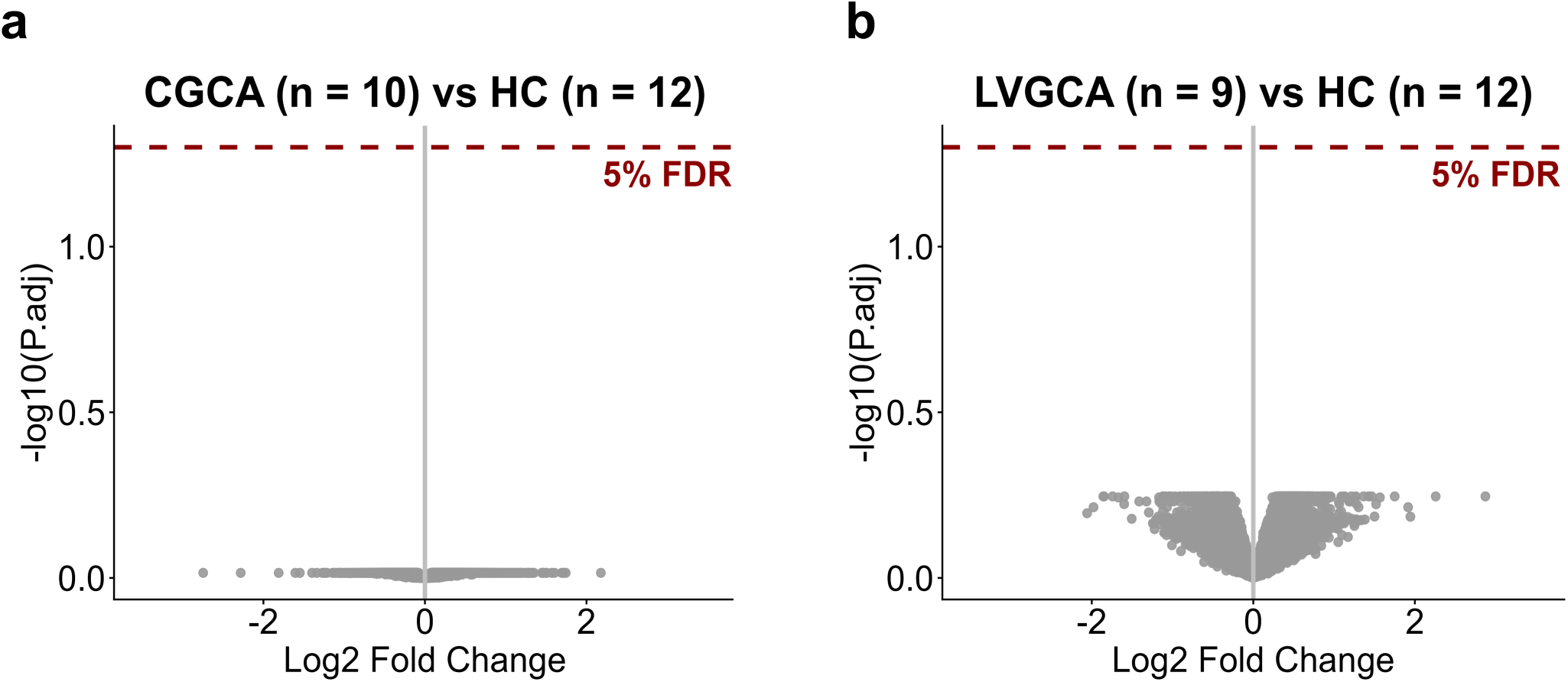
No evidence of autoantibodies in GCA subtypes. Volcano plots for stratified analyses comparing abundance of autoantibodies for each GCA subtype versus HCs. **a)** Cranial GCA (C-GCA) versus HCs **b)** Large vessel GCA (LV-GCA) vs HCs. Each point represents a protein target on the autoantibody array. Red horizontal dotted line represents significant threshold (5% FDR). Grey = non-significant. P.adj = P-value after Benjamini-Hochberg correction for multiple testing.

**Figure S7:**
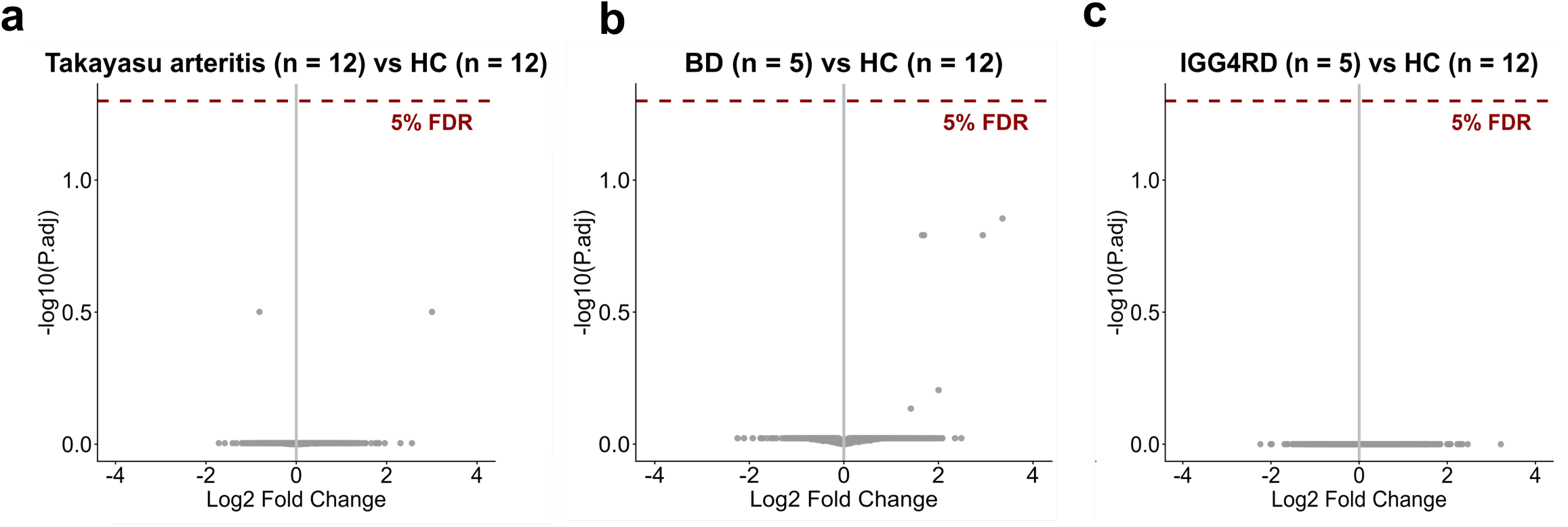
No evidence of autoantibodies in other immune-mediated diseases with vascular involvement. Volcano plot comparing abundance of autoantibodies against 15,286 autoantigens in other vascular inflammatory diseases vs healthy controls (HCs)**. a)** Takayasu arteritis vs HC **b)** Behcet’s disease (BD) vs HCs **c)** Immunoglobulin G4-related disease (IgG4-RD) vs HCs. Each point represents an antigenic target on the autoantibody array. Red horizontal dotted line represents significant threshold (5% FDR). Grey = non-significant. P.adj = P-value after Benjamini-Hochberg correction for multiple testing.

**Figure S8:**
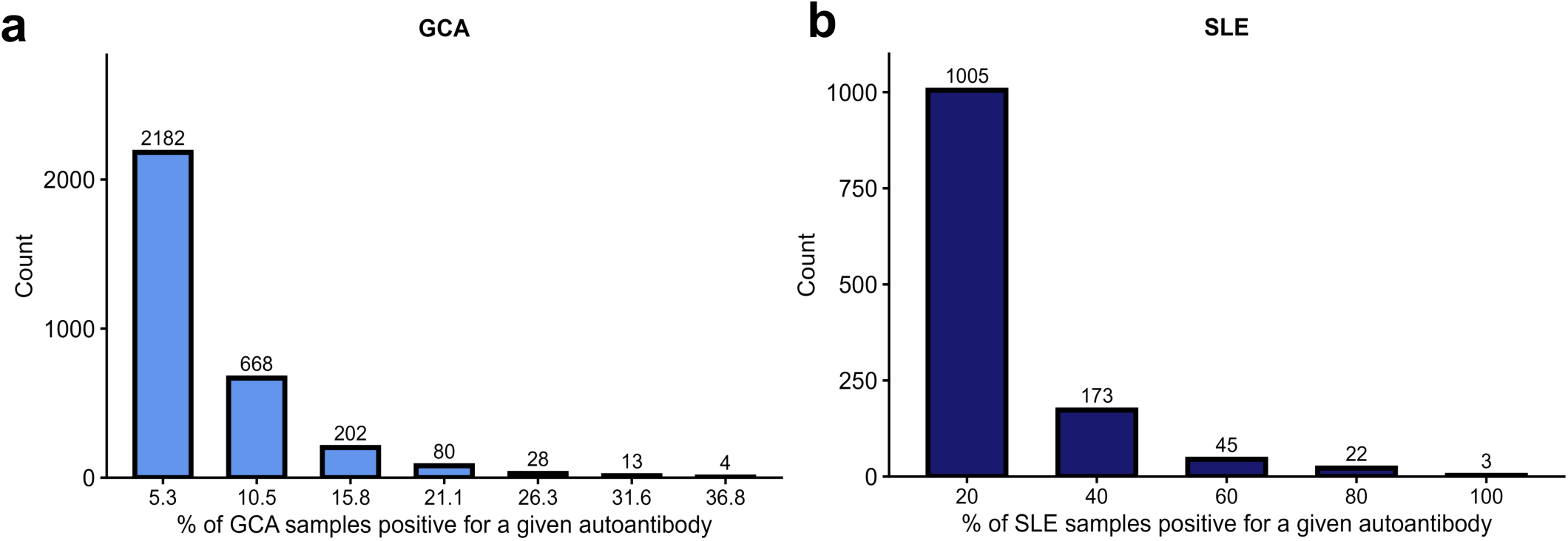
Distribution of positive autoantigen reactivity in GCA and SLE based on a healthy control-derived threshold. **a-b)** For each autoantigen assayed (15,286 targets in total), positivity was defined as a reactivity signal greater than the mean + 3 standard deviations of n=12 healthy control samples. The x-axis indicates the percentage of patients with positive reactivity to a given autoantigen. The y-axis indicates the number of autoantigens meeting this criterion within each prevalence bin. **a)** GCA (n=19) and **b)** SLE (n=5).

**Supplementary Data Tables 1-9** (accompanying Excel file).

<u>Key to Excel file:</u>

**Unique ID:** Unique identifier for each target

**Group:** Target type (full-length, protein isoform/peptide, or neoantigen)

**Name:** Gene symbol with unique suffix for each target

**Gene.ID:** National Center for Biotechnology Information (NCBI) gene ID

**HGNC.ID:** Human gene nomenclature committee (HGNC) gene ID

**Subject.accession.URL:** NCBI URL

**Translation_Emboss_AA:** Amino acid sequence for each target.

